# Stochastic simulation of successive waves of COVID-19 in the province of Barcelona

**DOI:** 10.1101/2021.12.02.21266468

**Authors:** M. Bosman, A. Esteve, L. Gabbanelli, X. Jordan, A. López-Gay, M. Manera, M. Martínez, P. Masjuan, Ll.M. Mir, J. Paradells, A. Pignatelli, I. Riu, V. Vitagliano

## Abstract

Analytic compartmental models are currently used in mathematical epidemiology to forecast the COVID-19 pandemic evolution and explore the impact of mitigation strategies. In general, such models treat the population as a single entity, losing the social, cultural and economical specifici- ties. We present a network model that uses socio-demographic datasets with the highest available granularity to predict the spread of COVID-19 in the province of Barcelona. The model is flexible enough to incorporate the effect of containment policies, such as lockdowns or the use of protec- tive masks, and can be easily adapted to future epidemics. We follow a stochastic approach that combines a compartmental model with detailed individual microdata from the population census, including social determinants and age-dependent strata, and time-dependent mobility information. We show that our model reproduces the dynamical features of the disease across two waves and demonstrate its capability to become a powerful tool for simulating epidemic events.

## Introduction

The COVID-19 outbreak struck the society like a tsunami, affecting citizens in many aspects, from mental health to economic well being. Communities all over the world had to modify habits and social contacts, in a common effort of erecting a barricade against the virus. Understanding the virus, its structure and its dynamics, is of paramount importance not only for the development of medical solutions (vaccines and specific pharmacological therapies) but also to define the social strategy of prophylaxis and prevent the expansion of the pathology.

The contagious spread of COVID-19 and the impact of mitigation strategies are currently the subject of many studies, all of them pursuing a better understanding of the variables at stake (it is worth citing the progenitor studies by Imperial College London ^1, 2^ and, in the particular case of Catalonia, those conducted by the Computational Biology and Complex Systems group at UPC ^3–5^; for further references see also the review by Estrada ^6^). This is a complex problem because it involves several factors acting simultaneously, with different weights and consequences, and both short- and long-term effects. What we know for certain is that the disease propagates mainly through social contacts ^7^. It becomes then crucial to understand the pattern of contacts of each individual – and how this is intertwined with demographic and social determinants – to comprehend the natural history of COVID-19, as well as the specific attributes (age, previous medical conditions, etc) that determine the outcome of the infection in each infected person ^8^. The frequencies and types of contacts are influenced by lockdown policies, while the probability to become infected depends on individual protection measures. Additionally, the probability for infected people to be diagnosed depends not only on the severity of the symptoms but also on testing policies in place. Global epidemiological indicators, like the incidence of the disease, hide a complex entanglement of highly diversified social and individual characteristics that perhaps can be ignored in a first approximation, but ends up being indispensable to model the epidemic.

Several approaches can be pursued. Monitoring indicators, like the daily number of positive tests ^3–5^, are useful to track the dynamical evolution of the outbreak in varying conditions and allow simple short-term extrapolations. These results, however, are very sensitive to initial conditions and need a continuous readjustment of inputs to be reliable in the long run. The exponential amplification of small perturbations in the initial data, typical of epidemics, makes the outcome of an outbreak intrinsically unpredictable ^9^. As for similar forecasting models, epidemic models cannot return exact predictions; epidemic models can, at most, analyse the likelihoods of different scenarios ^10^.

In this context, simulation tools turn out to be decisive to explore *in silico* the phase space of all the known variables and to unravel those steps in the contagion whose interdiction would prevent the epidemic progression (for example, by government policies regarding the introduction of tracking software, quarantine of exposed individuals, selective lockdown or mandatory use of protective masks in certain conditions). The purpose of such tools is thus threefold: first, they are *monitoring* tools, able to catch the evolution of the disease; second, they are *forecasting* tools, used to identify the conditions that could favour an outbreak; finally, they are *prevention* tools that can provide health authorities with studies on the effectiveness of containment measures. Analytical models as simulation tools usually treat the population as an aggregated body without taking into account its spatial variability or its social and demographic diversity. Some relevant features of COVID-19 can still be inferred from more sophisticated mathematical models (for Catalonia, for example, this has been done following a microscopic Markov chain approach applied to an age-stratified meta-population model ^11^). The loss of granularity, however, precludes the design of fine-tuned policies, which in turn renders the decision-making process not fully adequate in all aspects and for all groups, as proved not long ago for the case of H5N1 influenza ^12^.

In this work we present a different approach, a simulation tool that follows the daily contact network history of each individual in a largely populated area, the province of Barcelona. A precise knowledge of the morphology of these networks has been shown to be of the greatest importance to capture the salient characteristics of the outbreak ^13, 14^. For each individual, we consider their corresponding microenvironment: home location and the co-residence structure, the employment situation and the mobility routine with its resulting pattern of contacts. Using such information, we have developed a stochastic compartmental model that includes both population and epidemiological data. On the one hand, population data consist of all information about contact networks and mixing patterns (the “where“ and “how“ people live and move), based on the following external inputs:

i. Socio-demographic individual microdata from the latest census data provided by the Spanish National Statistics Institute ^15^.
ii. Contacts matrices for densely populated (and country-dependent) environments ^16–18^.
iii. Mobility data, reconstructed from information supplied by mobile network operators ^19, 20^.

On the other hand, epidemiological data ^8^ contain information related to the disease itself, its dynamics, and the different phases of an infectious process (exposure to the virus, infection, diagnosis and recovery). In this regard, we use a compartmental model, where the entire population is classified into five states: “susceptible”, “exposed”, “infected”, “diagnosed” and “recovered” (see **Methods**).

Our simulation tool calculates the daily probabilities for each individual of becoming infected at home, at work or school, during further social contacts (generically labelled as “Community”) and in public transport. Age, co-residence patterns, work activity, local mobility and the usage of public transportation are used to estimate the contacts network of each individual. Measures modifying the network and the frequency of the contacts (lockdown, teleworking, etc) are also taken into account, as well as the introduction of protective masks that modify viral load transmission (see **Methods**).

## Results

Our goal is to simulate the COVID-19 outbreak within the population of the province of Barcelona (5.5 million inhabitants) and reproduce the first two epidemic waves experienced in the region during February-June and July-December 2020 for a total of 300 days. We assume the existence of an initial set of about 50 exposed people (0.001% of the population), randomly distributed across the region. The actual location of people changes throughout the day and throughout the week: the weekdays of workers and students, for example, are divided into three 8-hour fractions according to a given pattern (home *⇒* job (or school) *⇒* other activities). The pattern of allowed locations and activities evolves according to lockdown and mobility restrictions promulgated during such 300 days.

The number of individuals in each epidemiological compartment predicted by the simulation as a function of time is shown in Figure 1. Without lockdown measures (Figure 1a), more than 90% of the total population of Barcelona and its province get affected by the disease in about 100 days. Taking into account lockdown measures (Figure 1b), the amount of infected people reduces to 300k in about the same period. In the second wave, continuing until the end of the year, the total number of infected people increases to 750k. Figure 2 presents the numbers of currently infected and newly diagnosed people predicted by the simulation as a function of time. The figure presents cumulative and age-dependent evolution curves. The subset of hosts of collective accommodation institutions (nursing homes) is shown separately. The cumulative curve shows a first wave characterised by a steep onset, interrupted by the strict lockdown measures established on March 16, 2020. The mobility restrictions induce the turn-over of the curve and its subsequent decay over a time scale related to the duration of the incubation and infectious phases of the disease. Contacts start increasing again during summer, leading to a second wave in autumn. Adults constitute the most relevant fraction of the infected compartment during both waves. The category of diagnosed is instead dominated by seniors in the first wave and adults in the second.

**Figure 1:**
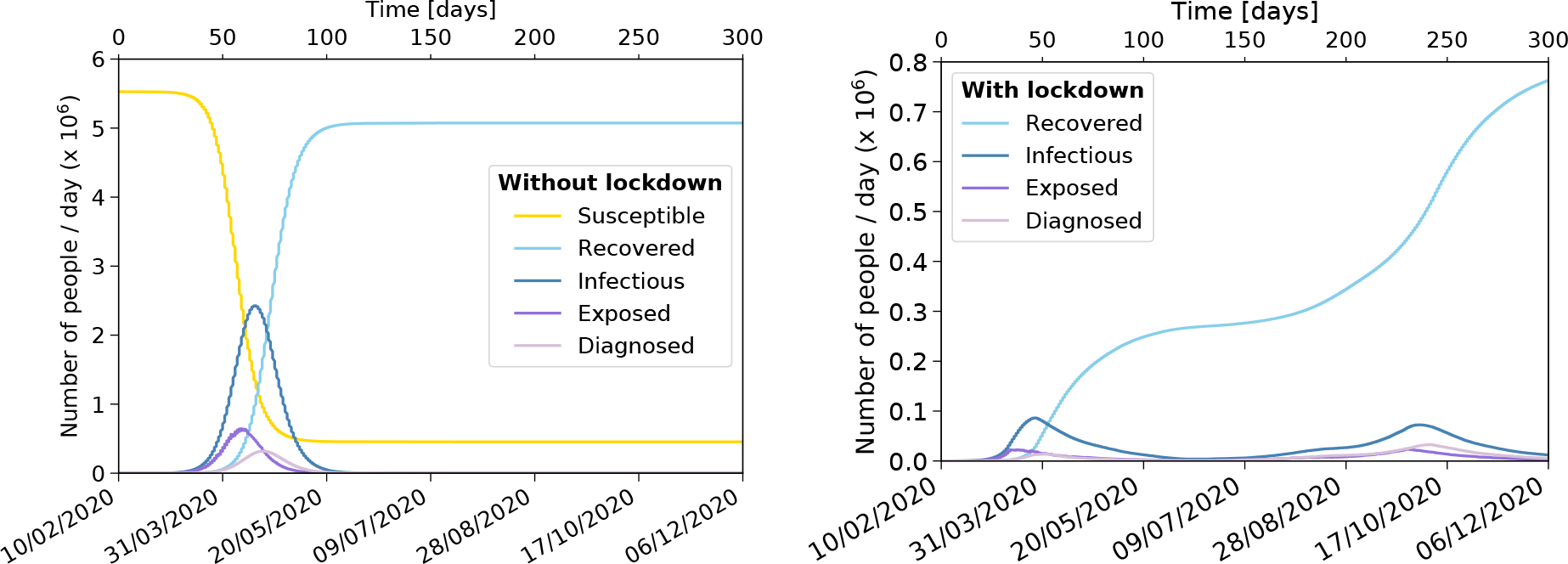
Evolution of the COVID-19 pandemic in the province of Barcelona. Number of susceptible, exposed, infected, diagnosed and recovered people as a function of time over a period of 300 days from February 10 to December 6, 2020. It has been assumed that 50 people, chosen at random, are exposed at time zero. **a** No lockdown measures. **b** Lockdown measures are applied [susceptible compartment is omitted].

**Figure 2:**
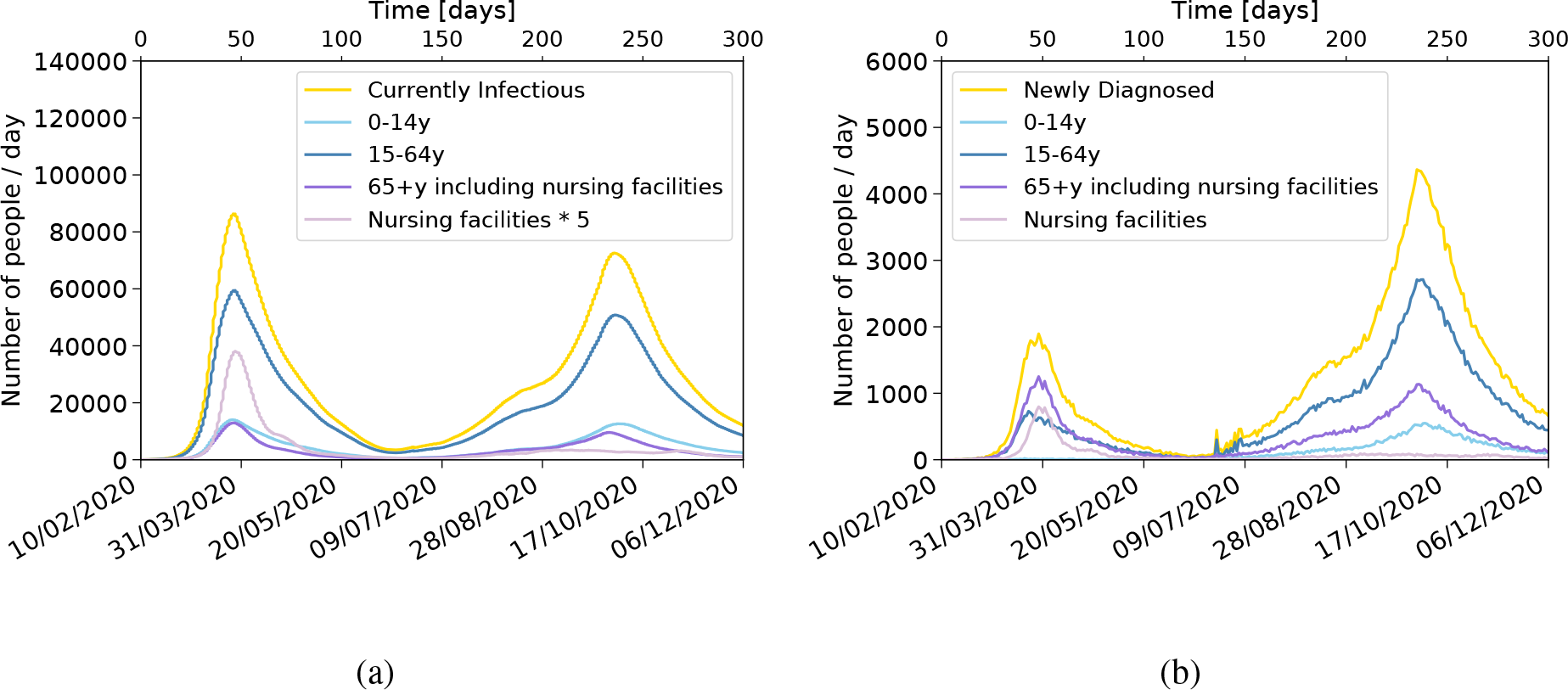
Time evolution of infected people. The number of infected people over a period of 300 days is shown split by age categories: less than 15 years old, from 15 to 64 years old and 65 years old and above (hosts in nursing facilities are shown separately). It has been assumed that 50 people, chosen at random, are exposed at time zero. **a** Total number of infected people in a given day. The number of people in nursing facilities is multiplied by five to make it more visible. **b** Newly diagnosed people every day. In this case, the number of people in nursing facilities is not multiplied by five.

Relevant results have been obtained by considering separately age-, social-activity- and region-dependence:

i. The number of diagnosed people, as shown in Figure 2b, can be compared with data pro- vided by the Catalan Health Service ^4^, as shown in Figure 3. The outcome of the simulation resembles quite closely the real picture emerged in these months for the number of positive PCR tests. Seniors in nursing facilities are notably more numerous than expected from their natural demographic fraction. This pattern results from the joint action of three factors. First, the probability of developing high viral load with strong symptoms increases significantly as a function of age starting at 65 years-old, while adults have a much lower probability to develop strong symptoms, and even less so children. Second, tightly knitted communities as those of nursing facilities favour multiple contacts. Third, during the first wave people with strong symptoms would reach the public health system faster and have a higher probability of being diagnosed. The features of the second wave with a peak in October are also qualitatively reproduced, with the spread of the disease picking up again during summer. In this case adults form the group dominating the diagnosed compartment, with children further contributing visibly. The special protection measures taken in nursing facilities (wearing protective masks) reduced contagion, while the amount of tests (and hence the probability to get diagnosed) increased and reached a larger part of the population. Our simulation shows that the unusual strength of the second wave has its root in extra occasional contacts taking place during the summer.
ii. We analysed under which circumstances people become infected either being at home, at work or school, or involved in social activities, such as shopping, leisure time, etc. Public transportation effects are taken into account: mostly during work time for those using mass transportation for their route to work, or during the weekends or the week for those citizens using group travel systems for other activities (see **Methods**). In our simulation, the amount of available environmental virus, or the viral load each individual is exposed to, is encoded in the *force of infection λ*, an aggregate parameter accounting for the probability to change the status of an individual from susceptible to exposed (see Equation 1 in **Methods**). The results in terms of the viral load are shown in Figure 4. This is an effective way to forecast the risk of contagion in the different social activities. The figure shows that in the early days before lockdown, the viral load exposition is roughly equal at home and at work or school, being slightly higher for community activities. However, when strong lockdown measures are applied on March 16, the contributions from work and community drop drastically and contagion at home becomes the most relevant contribution. The exposition to viral load in public transport or due to casual contacts during summer activities is relatively small, overall of the order of 1%, but is nevertheless a relevant component as it can bring the virus to households and work communities that would otherwise not be exposed to the infection.
iii. We investigate the spatial dependence within our simulation. Real data ^4^ and the corresponding simulation are shown in Figure 5a and Figure 5b, respectively, for three regions regrouped in three concentric areas (see **Supplementary Material**). At the center, the Barcelonès – 2.3 million inhabitants – exhibits a rather sharp increase of the number of diagnosed people in early July, and then a plateau until the development of the peak of the second wave. A first “ring” – 2.5 million inhabitants – shows a somewhat slower onset of the second wave with respect to the Barcelonès, with a similar clear peak for the second wave. A second “ring” – 0.8 million inhabitants –, exhibits a similar pattern with an even smaller onset during July and a less intense peak during October. The detailed features of the rise are related to the corresponding pattern of summer contacts. In our simulation we implemented a simple common pattern in all regions. The resulting rise is in average slower than in the data. On the other hand the relative size of the second peak appearing simultaneously in the three regions and related to resuming activities in September is reasonably well reproduced although it develops about two weeks earlier than in data.

**Figure 3:**
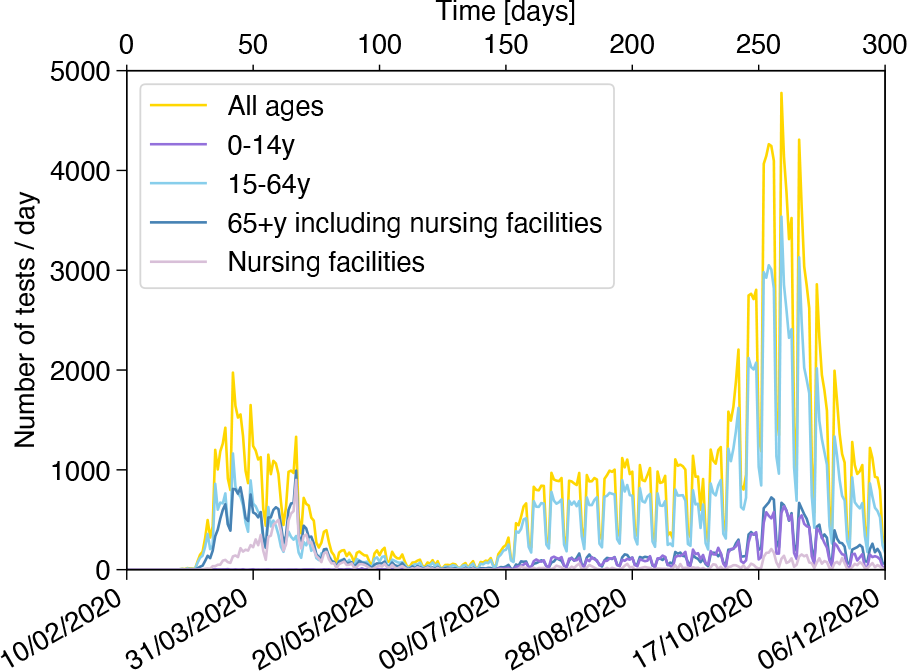
Number of positive PCR tests in the province of Barcelona. The number of positive PCR tests in the province of Barcelona for the period between February 10 and December 6, 2020 is shown separately for the different age segments of the population, including its sum.

**Figure 4:**
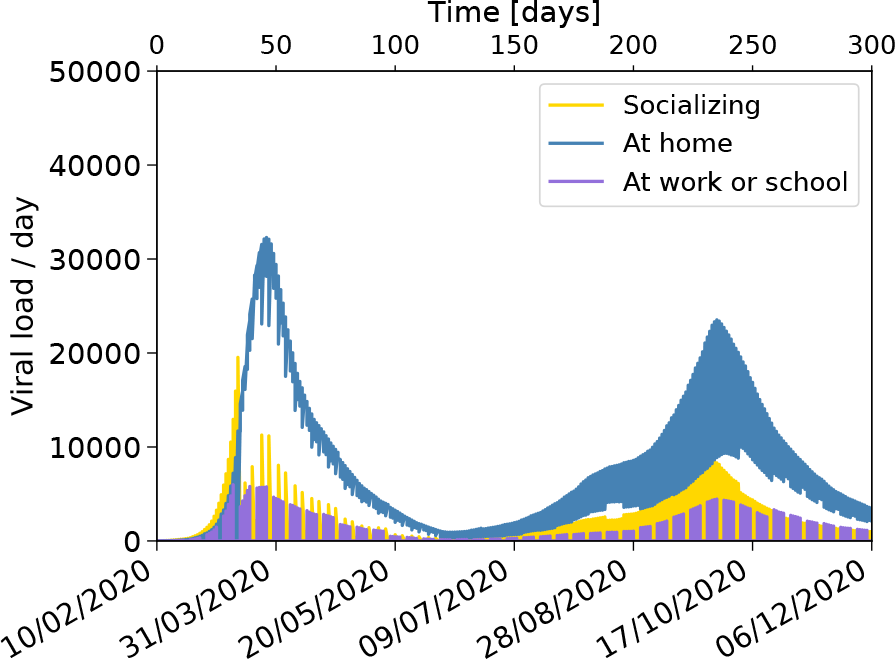
Total viral load evolution. The evolution of the main components of the viral load to which the population is exposed is shown separately for time spent at home, at work or school, or in social/community interaction. It has been assumed that 50 people, chosen at random, are exposed on February 10. The time evolution is shown for 300 days from February 10 to December 6, 2020.

**Figure 5:**
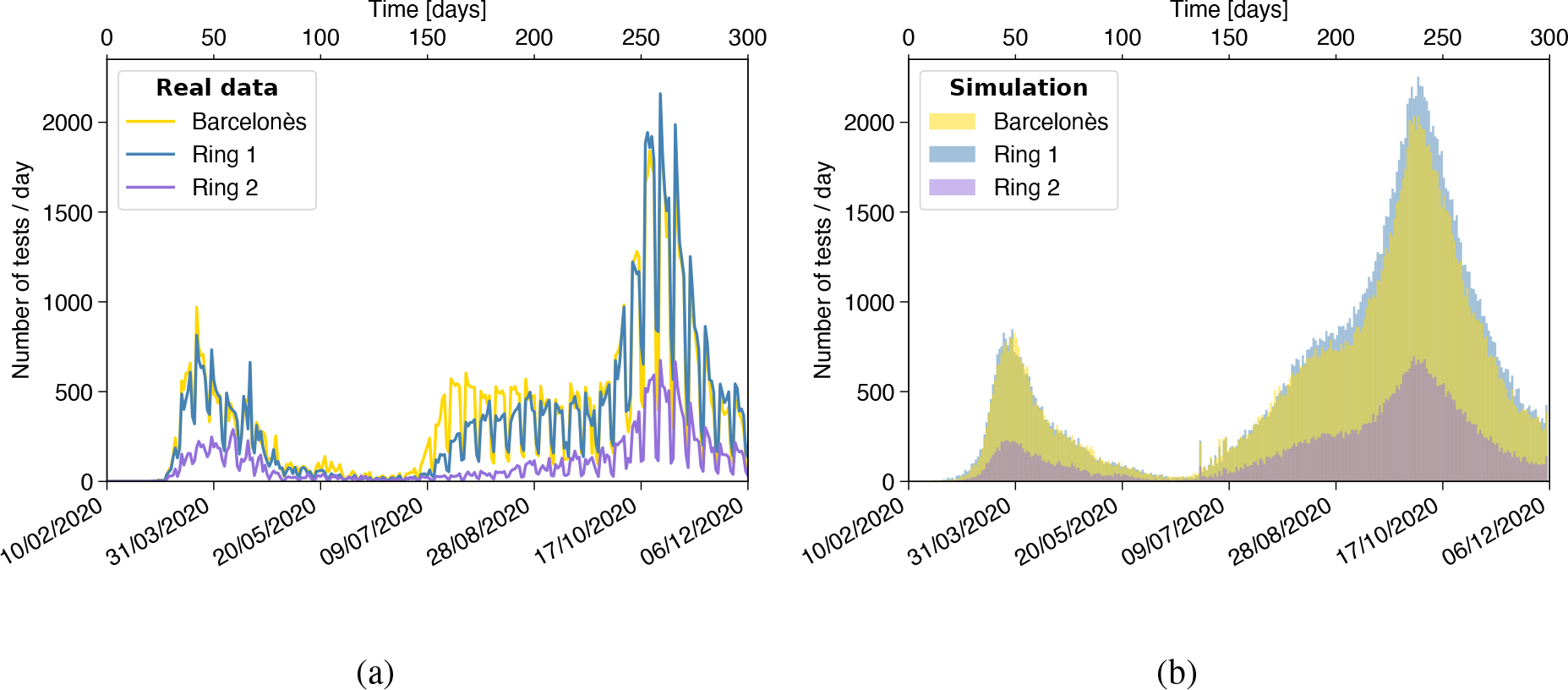
Number of positive PCR tests in the province of Barcelona. The results are shown separately for the Barcelonès county, and the counties surrounding it for the first and the second ring. **a** Real data. **b** Simulation.

## Discussion

The first and second waves are of quite different nature ^21^. The first one is the result of the initially free propagation of the virus, later controlled by the application of strict lockdown measures. Diagnostics were strongly related to the appearance of symptoms. During the second wave, mobility restrictions were alternatively relaxed and tightened depending on the epidemiological situation. The use of masks proved to be essential, as well as the increase of the efficiency of diagnosis of asymptomatic individuals and the corresponding quarantine measures.

The simulation tool presented here is able to reproduce the main features of the first and second waves of the COVID-19 outbreak in Barcelona and its province. The detailed description of two extremely heterogeneous categories of data, the sociodemographic and epidemiological ones, is a crucial ingredient. The sociodemographic individual microdata, convoluted with mobility information, are the base for modelling contacts and are necessary in order to correctly reproduce the expansion of the disease governed by the different components of the parameter *λ* (Figure 4). Indeed, since the contribution coming from the viral load components linked to work- and community-activities is drastically reduced as soon as lockdown measures prevail, co-residence contagion becomes the dominant factor, making it very valuable to use real census data, as done in our study. The incubation time, contagiousness period, and age-dependent strength of viral shedding are all important in order to predict the time profile of the viral shedding of an individual and the overall reaction time to lockdown measures.

Many ingredients are intrinsic to the population and do not change as a function of time, while others do vary. An important one is the probability of being diagnosed, which was initially linked to the appearance of strong symptoms, but later extended to people with less or no symptoms due to more awareness, contact-tracing and mass testing campaigns. As a result, the probability of detection in adults and children increased, reversing the proportions of diagnosed people by age-strata. The lockdown measures were continuously adjusted by the authorities to maintain a delicate equilibrium between the contention of the virus and sustaining the economic and educational activities. Mobile phone data, clearly correlated to these measures (see **Methods**) and available separately for work activities and leisure, are used to predict the time dependence of the level of contacts. The simulation also needs to take into account the increased number of contacts taking place during the summer period in order to generate the conditions for the surge of a second wave.

In conclusion, our simulation tool leads to a deep understanding of COVID-19 dynamics in the province of Barcelona. It is able to reproduce the very different characteristics of the two waves, the underlying dynamics of the second one being much more complex. The stochastic approach allows an easy implementation of the many relevant determinants of the problem with the maximum available granularity, including time dependence. The main advantage of the tool is its capacity to study the relative impact of individual factors and to identify the most critical ones (see **Methods**). The default configuration does reproduce the relevant features of the data, giving confidence in the prediction of relative variations. More detailed calibration and validation with higher granularity data over longer period of time should bring further insight and precision. The entire machinery can be easily adapted to study the epidemiological picture of any other region for which individual socio-demographic microdata are available. We can also implement the clinical history of hospitalised patients. In the constant stream of new information about the disease, further data regarding vaccination campaigns, coexistence of different variants of the virus, impact of super-spreaders, effect of local small outbreaks, etc., could be easily implemented without having to change the structure of the simulation.

## Methods

The simulation described in this work reproduces the first and second wave of the COVID-19 outbreak in the province of Barcelona using both epidemiological data and population statistics, linking the “who”, “where”, “how” and “when” with the characteristics of the virus itself. In this section we detail the different aspects of the calculation. In particular, we discuss: the underlying compartmental model; the data used to describe the population; the description of contacts between individuals; how mobility data are processed and used to simulate the impact of lockdown measures; the implementation of other mitigation strategies (such as the use of protective masks); how the model is calibrated with the data; and finally, a description of the sensitivity of the simulation outcome to some of the most relevant parameters. The latter gives insight on the uncertainty of the prediction related to the limited knowledge of the parameters and the probabilistic nature of the problem.

### SEIDR model

We consider a model where the population is divided into five compartments: “susceptible”, “exposed”, “infected”, “diagnosed” and “recovered”, as shown schematically in Figure 6 (note that in our model we do not include traditional vital dynamics: births and deaths). When susceptible individuals enter in contact with infected persons, their state may change to exposed according to the probability

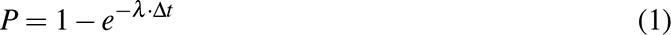

**Figure 6:**
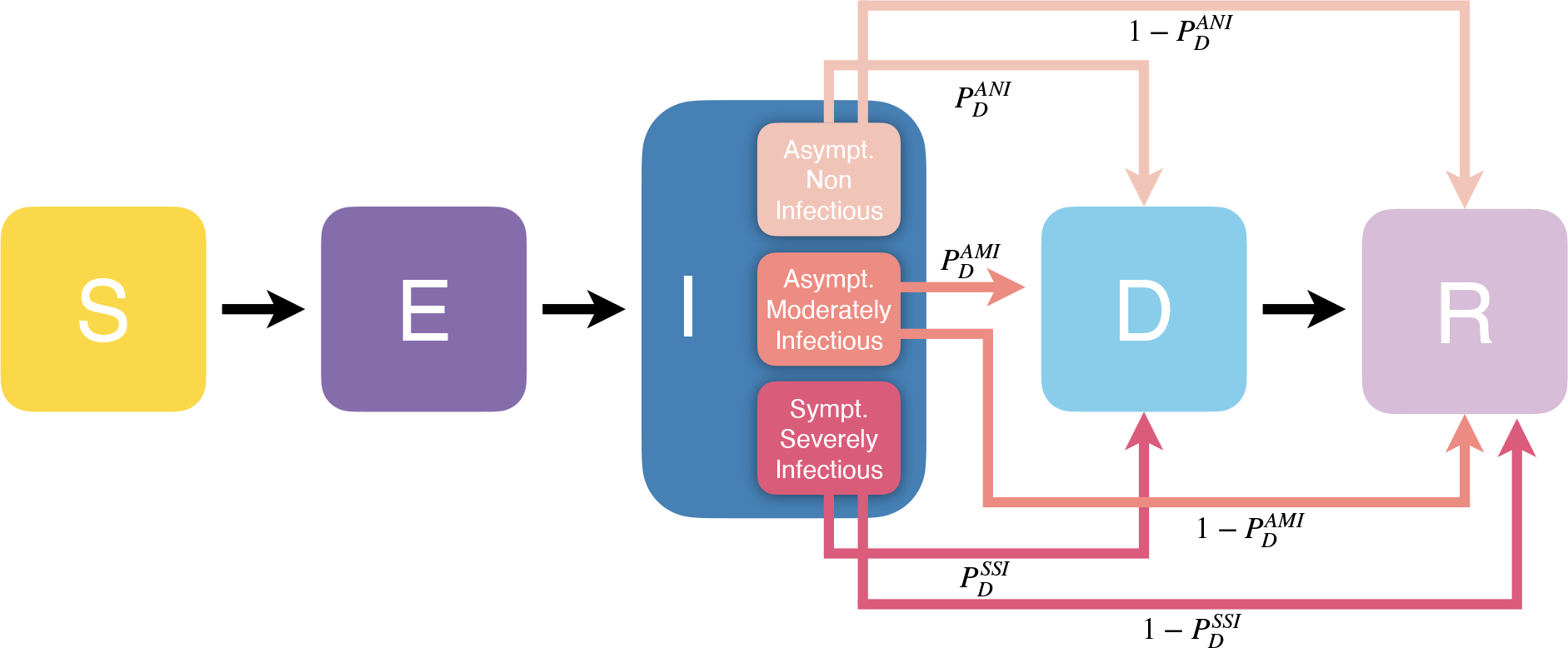
Transition diagram for our SEIDR model. When becoming infected, individuals are assigned a sub- compartment of disease characteristics with different symptomatology and infectiousness according to their age strata (0-14y; 15-64y; 65+y): asymptomatic non infectious (ANI), asymptomatic moderately infectious (AMI) and symptomatic strongly infectious (SSI). In turn, individuals are assigned the specific probability of being diagnosed of their sub-compartment (*PD*) and conversely of transiting directly to the recovered compartment 1 *− PD*.

where the force of infection *λ* is proportional to the total viral load to which the individual is If exposed, individuals are assigned a given set of variables to describe the evolution of the disease in their specific case. Every exposed person will eventually transit to the infected state. Epidemiological data are available for the incubation time defined as the time between infection and onset of symptoms ^22^. These data are described by a Gamma distribution with average of *µ* = 4.58 days and standard deviation *σ* = 3.24 days. The most probable value for incubation time is 3 days and the maximum allocated incubation time is 14 days. Epidemiological data show that the infectiousness process (shedding of virus) starts about two days before the onset of symptoms ^23^. The beginning of that process defines the transition from the susceptible to the infected state in the simulation and the beginning of the viral load shedding, whose time dependence is described by a Gamma function with average *µ* = 2.5 days and *σ* = 1.5 days. The maximum emission takes place after about 1.3 days. So, people shed a significant part of their total emission of viral load before the appearance of symptoms.

Further characteristics of the disease are set in the simulation according to the results of studies of epidemiological data from the beginning of the epidemics ^22–28^. Each member of the infected compartment is assigned one of three classes of symptoms and infec- tiousness: asymptomatic non-infectious (ANI), asymptomatic moderately infectious (AMI) and symptomatic strongly infectious (SSI) (Figure 6). The probability of belonging to such classes is strongly age-dependent: older people have a higher probability to develop symptoms and be more infectious, while children are more likely to be asymptomatic non-infectious. In addition, we assume a relation between the intensity of symptoms and infectiousness: infectious people with symptoms emit about twice as much viral load as asymptomatic infectious. In turn, each category has a specific associated probability of being diagnosed, which varied along the year. During the first wave, PCR tests were done mainly for people with strong symptoms, while during the second wave they would include close contacts of identified cases and random mass screenings. This is reflected in the setting of probabilities of detection. In the first wave *P*^SSI^_D_ = 1, while *P*^ANI^_D_ and *P*^ANI^_D_ = 0. In the second wave *P*^SSI^_D_ = 1, while *P*^ANI^_D_ = 0.4 and *P*^AMI^_D_ = 0.5. As a result, the probability to be diagnosed for the age strata 65+y is 70% in the first wave and goes up to 85% in the second wave, for the age strata 15-64y the probability changes from 15% to 55%, and for the age strata 0-14y from 1% up to 30%. In the simulation, the transition takes place in eight steps between June 25 and July 7, causing some discontinuities in the daily number of newly diagnosed people as seen in Figure 2b. People diagnosed are informed of the result after a time calculated according to a Poisson distribution with an average of six days since the start of infectiousness, a minimum of three and a maximum of 14 days ^27^. In the simulation diagnosed individuals are immediately put in effective quarantine and are thus no more contagious. All infected and diagnosed people will eventually move to the recovered state, at the latest 14 days after the start of infectiousness ^29^. Hospitalisation, admission to intensive care and possible death have not been implemented so far. Recovered people are considered immune.

### Population data

To simulate the history of each of the 5.5 million citizens of the Catalan province of Barcelona, we create a directory, based on the most updated census (“Cens de població i habi- tatges”) ^15^, with an entry for every individual. The original data set consists of approximately 400,000 people, each carrying an effective weight to account for the whole population. Each el- ement is characterised by a household identifier, age, gender and further relevant characteristics (e.g., occupation, location of the workplace, commuting time - if any -, commuting means of transport - if any -, etc.). The province of Barcelona is divided into 83 areas: notably, the city of Barcelona is divided into 51 neighbourhoods, while the rest of the province is split into 32 regions distributed in concentric areas around Barcelona city (see **Supplementary Material**).

### Nursing facilities

During the first months of the health emergency, the COVID-19 disease caused a high number of deaths among the elderly. In particular, the long-term care facilities suffered from an unfortunate notoriety due to the high mortality rate of their residents compared to people of the same age group living in family dwellings. The census we employed to reconstruct the population does not take into account people aged 65 years or over who reside in nursing facilities. In order to include this sector of the population, we used the list of the 756 official long-term care facilities established in 2019, along with details regarding the available places therein30. The relative age structure of the elderly population living in these facilities is almost symmetrically inverse to that of the elderly population living in family dwellings^31^: the oldest among the elderly live mainly in institutions, while the youngest live in private homes. Given the average occupancy level of nursing facilities at 86%^32^ and the location of each of the facilities, we reconstruct the profile for the estimated population of 40,000 individuals hosted in nursing homes. The ages of each of the individuals are randomly assigned according to the overall age structure.

### Description of activities and contacts

The history of activities of every individual is simulated as a function of time. The time is organised in weeks, with five labour days and two weekend days. A day is subdivided into three 8-hour intervals. The three daily time intervals correspond to time spent: i) at home, ii) at work/school for workers and pupils (or at home for the rest), and iii) in generic *social activities*. During the weekend, people are either at home or involved in social activities. The weekly pattern of occupations, together with the granularity of 8-hour interval, is customizable. This pattern is modified with time according to lockdown measures (e.g., closure of schools or promotion of teleworking). We define five categories of contacts:

i. Home: The home contacts include the explicit group of individuals sharing the same household or nursing facility. The average number of home contacts per person is 2.73.
ii. Work and School: We assume closed groups whose sizes follow a Poisson distribution with mean 10 for work, and fixed sizes for school classes varying from seven for 0-year-old to 28 above 12-year-old ^33^. In the simulation all workers and students are assigned a company or a school class, which is constant for the whole simulation period. Contacts happening in these reduced groups are stable and long-lasting. In both cases, we use the census information on both the proximity of the workplace and commuting time to simulate a geographical distribution of companies and schools. There are 2.2M workers and 950k school pupils. On average, the resulting number of contacts per person at work or at school is 13.6.
iii. Community: Here we include social contacts (other than in the family or occupational bubbles) that we generically label as “Community” contacts. The number of assigned contacts is distributed as a Gaussian with means taken from the largest and most up-to-date synthetic contact matrices set ^16^. The average number of contacts is age-dependent, but not the age profile of these contacts. Contacts are randomly assigned within the same region and are again assumed to be stable contacts but not of the “bubble” type: indeed, each element *A* has its own fixed “Community” network *C* (the “friends circle of *A*”), however, an element *B ∈ C* has a “Community” network *C I* that does not necessarily coincide with that of *A*. On average, such contacts sum up to 4.8 per person.
iv. Public transport: A fraction of people use public transportation to commute and a further fraction for community activities. This may lead to additional occasional contacts beyond the network of stable contacts described above. Using the most recent survey on the use of means of transport in the province of Barcelona ^17^, we estimate that 20% of schoolchildren plus 30% of workers use public transport on their way to school and workplace. We further estimate that 10% of the total population uses public transport for other personal reasons. We assume an effective number of about one occasional such contact per round trip and that the users all carry the average viral load of the population using public transport to go to work/school, or for community activities, depending on the case.
v. Occasional contacts during community activities:

Finally, there may be further contacts of occasional nature, similar to the case of the public transport. We assume that such contacts took place during the summer 2020 as a result of the relaxation of lockdown measures coinciding with typical summer activities in crowded outdoor settings (traditional festivals, cultural events, beach tourism). Some of these activities included also the additional presence of travellers from outside the province, modifying the effective population^34^. In the simulation, the number of such contacts is set to one per person with people carrying a viral load equal to the average of the full population.

### Mobility data

To control the spread of the disease, authorities imposed lockdown measures restricting the mobility, starting from March 16, 2020 and lasting for the rest of the year. We use the mobility data provided by the Instituto Nacional de Estadística (INE), extracted from the analysis of the position of more than 80% of the mobile phones by the three main Spanish mobile phone operators ^19^. Only phones with Spanish numbers are considered in the study. The INE aggregated these data into subgroups of population that remain, enter or leave a set of “mobility areas” ^20^. Data are provided for the year 2020, starting from March 16, both for weekdays and weekends. They are normalised to reference data from November 2019. We use the average data of the full Barcelona province as an indicator of work mobility and leisure mobility. The normalised intensity of mobile phones traffic is shown in Figure 7.

**Figure 7:**
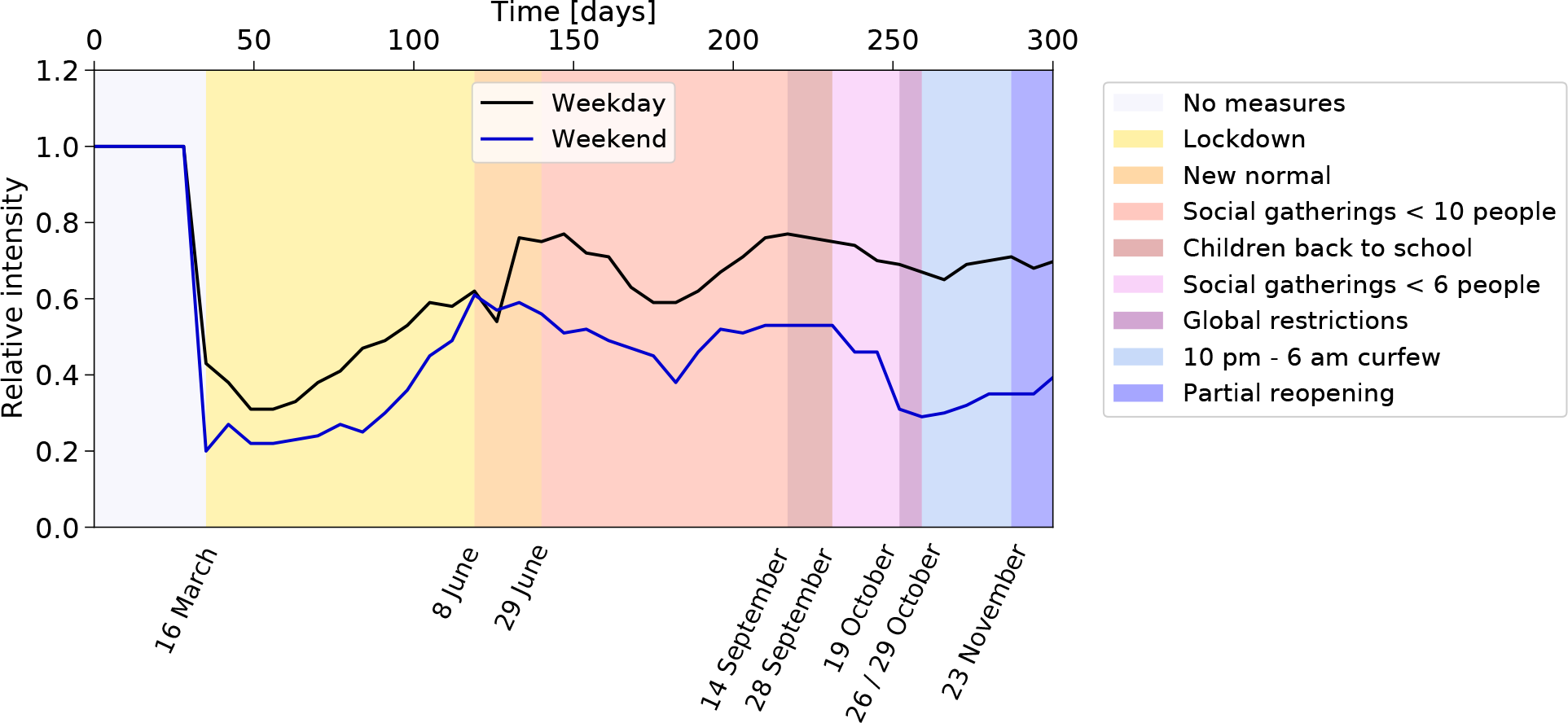
Relative intensity of mobile phones traffic and measures affecting work and community activities. Ratio between the intensity of mobile phones traffic during the period covered by the simulation, and for a reference week in November 2019. The pattern is influenced by the several containment measures adopted along the year. On March 16, 2020 the Spanish government decreed a state of alarm imposing of a succession of lockdown measures starting on the same day and lasting until June 8. The number of people in gatherings was limited to 10 and 6 persons on June 29 and September 26, respectively. Teleworking was encouraged, shops’ and cultural events’ capacity was reduced to 30% and 50%, respectively. Among other containment measures enforced on October 19, restaurants were closed, and university lectures made virtual.

### Lockdown measures

To simulate the impact of the lockdown measures ^35^, we combine two methods. Measures like school closure are directly implemented by changing the configuration of the activity in the time intervals. For example, pupils stay at home instead of going to school, and do not use public transport. For the reduction of work or community contacts, we rely on the time evolution of the mobility, as justified by the correlation between mobility changes and confinement measures observed in Figure 7. There is a a substantial dip in mid-March when the first lockdown was implemented, after which the mobility slowly increases. A smaller effect is observed around mid-August (possibly related to holidays), and a more pronounced one later towards the end of October associated with a tightening of the measures. Such correlation was also observed in other regions across Europe ^36^. In the simulation, we use the reduction of mobility to calculate the fraction of citizens teleworking or staying at home instead of participating to social activities.

### Further mitigation strategies

The use of protective masks was initially deterred (mostly due to the attempt of diverting the available masks to the healthcare providers), later encouraged and finally imposed. Studies show that wearing masks reduces virus transmission, although the extent of such reduction is difficult to quantify precisely ^37, 38^. The effect of wearing a mask is simulated by introducing a factor reducing the viral load transmission at work and in community activities and set to roughly estimated values, 0.7 when it was encouraged and 0.35 when imposed. We assume that people in nursing homes started wearing masks inside the facilities as a measure of precaution. We consider that part of the population, mostly young adults, relaxed the mask-wearing discipline towards the end of the year. In the simulation, we limit the reduction of viral load transmission from 0.35 to 0.85 for 13 to 35 years old people, from the end of October onward.

### Calibration procedure

The simulation counts with a total of 64 parameters to characterise the disease (26), the contacts (32) and the lockdown and self-protection measures (6) (see **Supplementary Material**). Most of them are set according to external information. Out of the 64 parameters that control the simulation, only 8 are adjusted with data as described below. First, we need to calibrate the scale factor converting the viral load calculated in arbitrary units into the force of infection leading to the correct probability of being infected. This probability governs the time needed to reach the onset of the explosive contagion regime and its growth rate. Second, for each specific value of the calibration parameter, we adjust the starting time of the lockdown measures in the simulation in order to approximately reproduce the rate of 1500 diagnosed people per day observed in the data at the peak of the first wave, as shown in Figure 3. We check that the growth rate is well reproduced: it should take about 10 days to go from 15% to 85% of the peak value. After adjusting the time, we see that the initial 50 infections happened five weeks before the start of lockdown measures, on February 10. Although the arrival time of the first infectious people in Catalonia is not precisely known, such date seems reasonable in view of the international context of the development of the disease. Third, the three categories of infectiousness need to be assigned a probability to be diagnosed for the first and the second wave. The probability of the strongly symptomatic to be diagnosed is set to one in both cases. The probability to be diagnosed for asymptomatic non-infectious and infectious was initially very low and increased significantly during the second wave. These two parameters are set to zero in the first wave and have been adjusted in order to approximately reproduce the observed proportion of diagnosed people by age-categories in the second wave (Figure 3). Fourth, additional summer contacts are needed in order to increase the risk of regrowth and facilitate the appearance of a second wave. The four corresponding parameters (number of contacts, segment of population affected, and starting and ending dates) have been adjusted accordingly, in order to also respect the observed proportions of diagnosed people in the different regions (Figure 5a).

### Sensitivity to Parameters

We studied the sensitivity of the simulation to the parameters and identified the most critical ones. This is relevant to understand the weakest points of the mitigation strategies. It also indicates what should be better known for an improved precision of the simulation.

First we report on the impact of parameters governing the response of individuals to the disease.

i. If we reduce the overall scale of infectiousness by a factor of two, it takes twice as much time to reach the explosive contagion regime.
ii. Reducing by a factor of two the probability to be diagnosed in the asymptomatic non-infectious and asymptomatic infectious categories during the second wave significantly reduces the number of people being diagnosed, and subsequently put in quarantine. As a consequence a factor of 2.5 more people become infected and the number of people diagnosed increases by 30%.
iii. The time it takes for people to get diagnosed is drawn from a Poisson distribution with mean six ^27^. A minimum of three days is set for the notification of the diagnosis. Removing the minimum does not have much impact, but setting it to large values does. If it is set to six days, people are put later in quarantine and more contagions can take place. The number of diagnosed people in the second wave doubles at the same time as the delay in the notification delays the peak by about a week.
iv. The effectiveness of the mask is important in the surge and strength of the second wave. The simulation shows that without that protection a second wave would have quickly developed. Changing the nominal reduction of viral load transmission of 0.35 by *±*0.05 changes the number of diagnosed people by *±*50%. With much stronger reduction no second wave would develop.

Next we investigate the sensitivity to the pattern of contacts.

i. The size of the contact groups at work is given by a Poisson distribution with mean 10. Changing it to 5 (20) leads to a reduction (increase) of 15% of diagnosed people. The impact is smaller in the second wave due to teleworking.
ii. In addition to the 950k school pupils there are 95k university students. Since our census data did not provide detailed information for this group and there were essentially no presential classes at university during 2020 after the start of the first lockdown, we considered that this group neither studied, nor worked. To analyse the impact of this assumption, we aggregated them to the pupils, assuming an average size of closed groups of contacts of 15. They resumed presential classes in mid-September. As a result, the peak of the first wave increased by 25% without readjusting the time of lockdown, while the peak of the second went down by 25%.
iii. We studied the impact of making the number of community contacts dependent on the den- sity of the population. We tested a model where the proportionality factor varies from 0.6 to 2, between Berguedà and Barcelonès, respectively, the lowest and highest density regions. The average number of community contacts increases from 4.8 to 7.6. During the first wave, the total number of diagnosed increases by 30%. The overall effect is less noticeable during the second wave, but the number of diagnoses in the Barcelonès starts dominating, unlike what is observed in Figure 5a. Such a strong density dependence is thus disfavoured.
iv. The number of additional summer contacts is much more critical than the contacts in trans- port as they affect potentially a much more significant fraction of the population. Doubling the default level of one contact generates an earlier wave during the period where a rela- tively flat distribution is observed, while dividing by two reduces the level of diagnoses in early summer by a factor of six, and no second wave develops. We also tested the impact of limiting the extra contacts to the Barcelonès area rather than the whole province. However, this option does not reproduce well the sharing of diagnosed cases in the different regions (Figure 5a).
v. Another factor that may influence the evolution of the disease is the location of the initially infected people. In the default configuration, they are randomly distributed geographically in the province. We kept the same total number but randomly varied their location. In the first wave, this results in a variation of the onset time by *±*2.5 days. If we keep the starting date of lockdown fixed, the maximum of the first wave varies by about *±*30%. Indeed the starting time of the lockdown measures is quite critical, since one week delay typically results in a factor of three higher number of diagnosed people per day at the peak. The cases with upward fluctuations of the number of diagnosed people in the second wave are, in general, not correlated with the cases of upward fluctuations in the first wave. Only in about 1% of the cases, when the first wave developed very early, the number of infected people grew a factor of three more before the lockdown took effect (the date was kept fixed). These people eventually became immune, and later, no strong second wave was able to develop.

Finally, we studied variations in the description of mobility.

i. To test the relevance of mobility and its correlation with the time dependence of the number of diagnosed people, we artificially introduced an additional four weeks of flat level of mobility in July. This has a strongly correlated effect on the time-dependent pattern of the spread, showing that mobility is a crucial ingredient for describing the evolution of the pandemic.
ii. The mobility data came in two sets with different methods and normalisations. Increasing the relative normalisation of the second set (after mid-June) by 10% almost doubles the level of infected people and shifts the peak earlier by one week.

In summary, we have explored part of the phase space of the many parameters playing a role in the expansion of the virus. The most sensitive ones include the effect of the protective masks, the probability of diagnosis, the characteristics of seasonal (summer) contacts and mobility. In some cases, more precise external information can be gathered (e.g. likelihood of diagnostic), or the full granularity of the information could be used, leading to possible improvement (e.g. mobility per region). More complex are the cases of protective masks or summer contacts. Improvements could come from a more detailed description of the parameters together with a comparison with high granularity data in terms of age and location.

### Data availability

The census data used to build the complete description of the population of the Barcelona province are available from CED and the Instituto Nacional de Estadística (INE, Spanish National Institute of Statistics: 2011 Population and Housing Censuses). However, restrictions apply to the availability of these data, which were used under license for the current and previous studies, and so are not publicly available. The census data and the derived data describing the entire population of the Barcelona province are however available from the authors upon reasonable request and with permission of CED and INE. The data describing diagnosed people in 2020 in the province of Barcelona, aggregated by age or by county, are publicly available at the Agència de Qualitat i Avaluació Sanitàries de Catalunya (AQuAS) (https://aquas.gencat.cat/ca/actualitat/ultimes-dades-coronavirus/). The mobility data are publicly available from Instituto Nacional de Estadística (INE) https://www.ine.es/en/experimental/movilidad/experimental em en.htm.

### Code availability

The code used in the simulation is available upon request.

## Data Availability

All data produced in the present study are available upon reasonable request to the authors

## Acknowledgments

The authors affiliated to CED, IFAE and i2CAT acknowledge the support of the CERCA institution, Centres de Recerca de Catalunya. MB, XJ, LLM, MMar, PM, JP, IR acknowledge support from the grant 2020PANDE0180 of the programme PANDEMIES 2020, “Replegar-se per créixer: l’impacte de les pandèmies en un món sense fronteres visibles” of the Agència de Gestió d’Ajuts Universitaris i de Recerca of the Generalitat de Catalunya. LG thanks the funding from the European Union’s Horizon 2020 research and innovation programme under grant agreement ID 758145. AL acknowledges the support from the Talent Research Program (Universitat Autònoma de Barcelona). PM has received funding from the Spanish Ministry of Science and Innovation (PID2020-112965GB- I00/AEI/ 10.13039/501100011033), from the Agency for Management of University and Research Grants of the Government of Catalonia (project SGR 1069), and also received support from Ajuntament de Barcelona. MMan acknowledges support from Marie Sklodowska-Curie grant agreement ID 6655919. VV has been partially supported by the H2020 programme and by the Secretary of Universities and Research of the Government of Catalonia through a Marie Skłodowska-Curie COFUND fellowship – Beatriu de Pinós programme ID 801370.

## Author Contributions

MB led the conception of the project. MB, LLM, MMan, PM, VV contributed to the modelling design. MB, LG, XJ, AL, LLM, IR took care of data preparation. MB, LG, LLM, MMan, PM, AP, IR developed the code. MB, LLM, PM, VV wrote the manuscript. All authors contributed to the discussion and interpretation of the results, revised critically the draft and approved the final version of the manuscript.

## Supplementary material

### Parameters of the model

The model to simulate the spread of Covid-19 counts with a total of 64 parameters to characterize the disease (26), the contacts (32) and the lockdown and self-protection measures (6). Most of them are set according to external information. Out of the 64 parameters that control the simulation, only 8 are adjusted. Tables 1 to 7 describe the parameters, their settings and related references.

**Table 1:** Parameters of the model (part I).

| Category | Number | Parameters | Source | Adjusted |
| --- | --- | --- | --- | --- |
| Disease |  |  |  |  |
| Incubation time (Gamma distribution) | 2 | $\mu, \sigma = 4.58, 3.24$ days | Medical data <sup>1, 2, 3</sup> | No |
| Infectiousness time (Gamma distribution) | 2 | $\mu, \sigma = 2.5, 1.7$ days | Medical data <sup>1, 2, 3</sup> | No |
| Onset of infectiousness in relation to onset of symptoms | 1 | -2 days | Medical data <sup>2</sup> | No |
| Relative viral strength of asymptomatic, moderate and severely infectiousness | 3 | 0, 1, 2 | Medical data <sup>3</sup> | No |
| Viral strength to probability of infection conversion | 1 | 4 | Adjusted | Shape & size of 1<sup>st</sup> wave |
| Fraction in categories of viral strength (age dependent) | $2 \times 3$ | See table 3 | Medical data <sup>3</sup> | No |
| Probability to be diagnosed in first (second) wave | $3 \times 2$ | $P_D^{ANI} = 0 \quad (0.4)$<br>$P_D^{AMI} = 0 \quad (0.5)$<br>$P_D^{SSI} = 1 \quad (1)$ | Medical data <sup>4</sup> | Yes for $P_D^{ANI}$ & $P_D^{AMI}$ during 2<sup>nd</sup> wave |
| Diagnosis time (Poisson distribution) | 3 | $\mu$, minimum, maximum = 6, 3, 14 days | Medical data <sup>4</sup> | No |
| Recovery time | 1 | 14 days | Medical data <sup>5</sup> | No |
| Initially exposed | 1 | 0.001% | Estimated | No |
| Contact's model |  |  |  |  |
| Workplace size (Poisson distribution) | 1 | $\mu = 10$ | Estimated | No |
| Classroom size (fixed, age dependent) | 6 | See table 4 | Departament d'Educació, Gen-Cat <sup>6</sup> | No |
| Social contacts (Poisson distribution, age dependent) | 16 | See table 5 | Synthetic contact matrices <sup>7</sup> | No |
| Spurious summer contacts | 4 | See table 6 | Ajuntament de Barcelona <sup>8</sup> | Shape of 2<sup>nd</sup> wave |

**Table 2:** Parameters of the model (part II).

| Category | Number | Parameters | Source | Adjusted |
| --- | --- | --- | --- | --- |
| Use of public transportation |  |  |  |  |
| Fraction of workers (Work) | 1 | 30% | Autoritat del Transport Metropolità (ATM) <sup>9</sup> | No |
| Fraction of pupils (School) | 1 | 20% | ATM <sup>9</sup> | No |
| Fraction of population (social activities) | 1 | 10% | ATM <sup>9</sup> | No |
| Average number of contacts per round trip (work/school) (fixed number) | 1 | 1.0 | Estimated | No |
| Average number of contacts per round trip (social activities) (fixed number) | 1 | 1.1 | Estimated | No |
| Confinement |  |  |  |  |
| Date of 1st day of simulation | 1 | 34 days before March 16, 2020 starting date of first confinement | Diari Oficial de la Generalitat de Catalunya <sup>10</sup> | Shape of 1<sup>st</sup> wave |
| Effect of mask | 4 | See table 7 | Medical data <sup>11, 12</sup> | No |
| Inter-calibration of mobile data | 1 | 1 | Estudios de movilidad a partir de la telefonía móvil <sup>13</sup> | No |

**Table 3:** Fraction of asymptomatic, moderately and severely infectious people depending on the age.

| Age (years) | Asymptomatic | Moderately infectious | Severely infectious |
| --- | --- | --- | --- |
| < 15 | 0.69 | 0.30 | 0.01 |
| 15 – 64 | 0.40 | 0.45 | 0.15 |
| > 64 | 0.05 | 0.25 | 0.70 |

**Table 4:** Number of children in each age group.

| Age | Classrooms' size |
| --- | --- |
| 0 | 7 |
| 1 | 12 |
| 2 | 16 |
| 3 – 5 | 22 |
| 6 – 11 | 25 |
| 12 - 18 | 28 |

**Table 5:** Average number of stable social contacts depending on the age.

| Age | Mean |
| --- | --- |
| < 5 | 3.21 |
| 5 – 10 | 3.88 |
| 10 – 15 | 5.27 |
| 15 – 20 | 5.97 |
| 20 – 25 | 5.26 |
| 25 – 30 | 4.33 |
| 30 – 35 | 4.35 |
| 35 – 40 | 5.51 |
| 40 – 45 | 6.48 |
| 45 – 50 | 4.94 |
| 50 – 55 | 5.26 |
| 55 – 60 | 5.26 |
| 60 – 65 | 5.06 |
| 65 – 70 | 4.05 |
| 70 – 75 | 4.89 |
| > 75 | 2.95 |

**Table 6:** Parameters used to model additional summer contacts.

| Parameters | Value |
| --- | --- |
| Average number of additional contacts | 1.0 |
| Starting date | June 8, 2020 |
| Ending date | September 28, 2020 |
| Fraction of population involved | 100% |

**Table 7:**
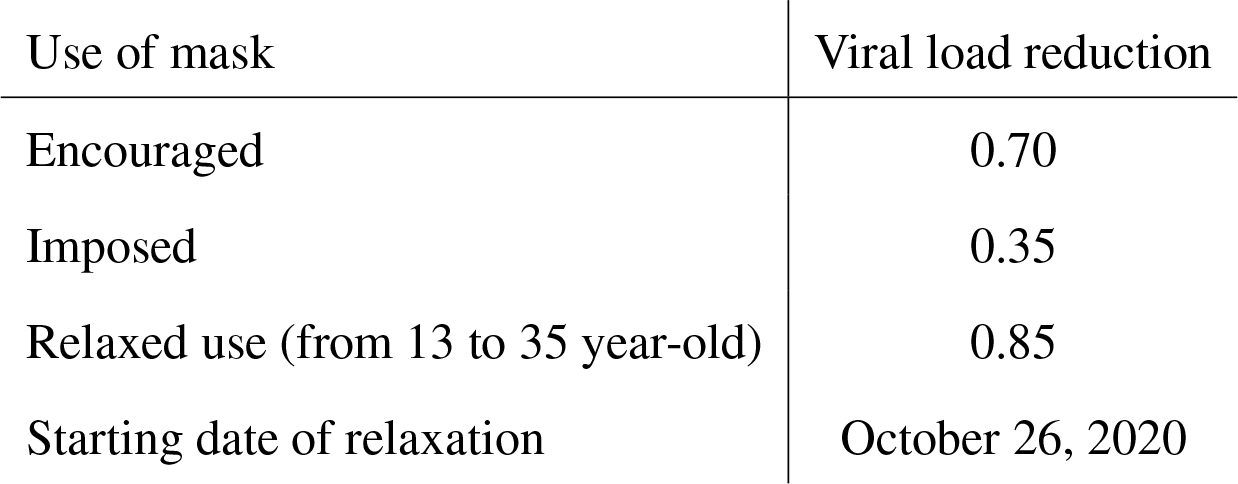
The effect of wearing a mask is simulated by introducing a factor that reduces the viral load transmission at work or school and in community activities. As of October 26, 2020, a relaxation of the discipline in the use of the masks by young adults is assumed.

| Use of mask | Viral load reduction |
| --- | --- |
| Encouraged | 0.70 |
| Imposed | 0.35 |
| Relaxed use (from 13 to 35 year-old) | 0.85 |
| Starting date of relaxation | October 26, 2020 |

### Statistics of the sample

Tables 8, 9 and 10 present the statistics of different subsets of the population.

**Table 8:** Number of people in each age category.

| Age (years) | Persons |
| --- | --- |
| < 15 | 861,266 |
| 15 – 64 | 3,704,595 |
| > 64 | 961,372 |
| → in nursing homes | 39,373 |
| Total | 5,527,233 |

**Table 9:** People working or going to school.

| Occupation | Persons |
| --- | --- |
| Workers | 2,247,353 |
| Schoolchildren (0 to 18 year-old) | 953,019 |
| Other | 2,326,861 |
| Total | 5,527,233 |

**Table 10:** Workplaces model assumed using the information available in the census file.

| Work location | Home region | Time to work | Assigned work region | Workers |
| --- | --- | --- | --- | --- |
| At home | Any | – | Home region | 197,827 |
| Own municipality | BCN | < 20' | Home neighbourhood | 190,316 |
|  |  | > 20' | Another BCN neigh. | 254,219 |
|  | Counties | Any | Home region | 448,229 |
| Another municipality | BCN | Any | Counties | 128,165 |
|  | Counties | < 45' | Home region | 659,138 |
|  |  | > 45' | Outside home region | 175,293 |
| Several municipalities | BCN | Any | Counties | 42,032 |
|  | Counties | Any | Home region | 144,856 |
| Abroad | Any | Any | Outside home region | 7,278 |

### Description of the province of Barcelona

In the simulation, the province of Barcelona is divided into 83 regions: notably, the city of Barcelona is divided into 51 neighbourhoods, while the rest of the province is split into 32 regions. Table 11 shows the number of inhabitants in the 32 regions. These regions are distributed within three concentric areas with the city of Barcelona at the center, as shown in Figure 1: the Barcelonès – the county including the cities of Barcelona, Badalona, L’Hospitalet de Llobregat, Sant Adrià de Besòs and Santa Coloma de Gramanet, for a total of 2.3 million inhabitants –, a first “ring”, which includes Baix Llobregat, Vallès Occidental, Vallès Oriental, and Maresme – 2.5 million inhabitants –, and a second “ring”, including Garraf, Alt Penedès, Anoia, Bages, Berguedà, and Osona – 0.8 million inhabitants –. Table 12 shows the number of inhabitants in the 51 neighbourhoods of Barcelona, for a total of 1.6 million people.

**Figure 1:**
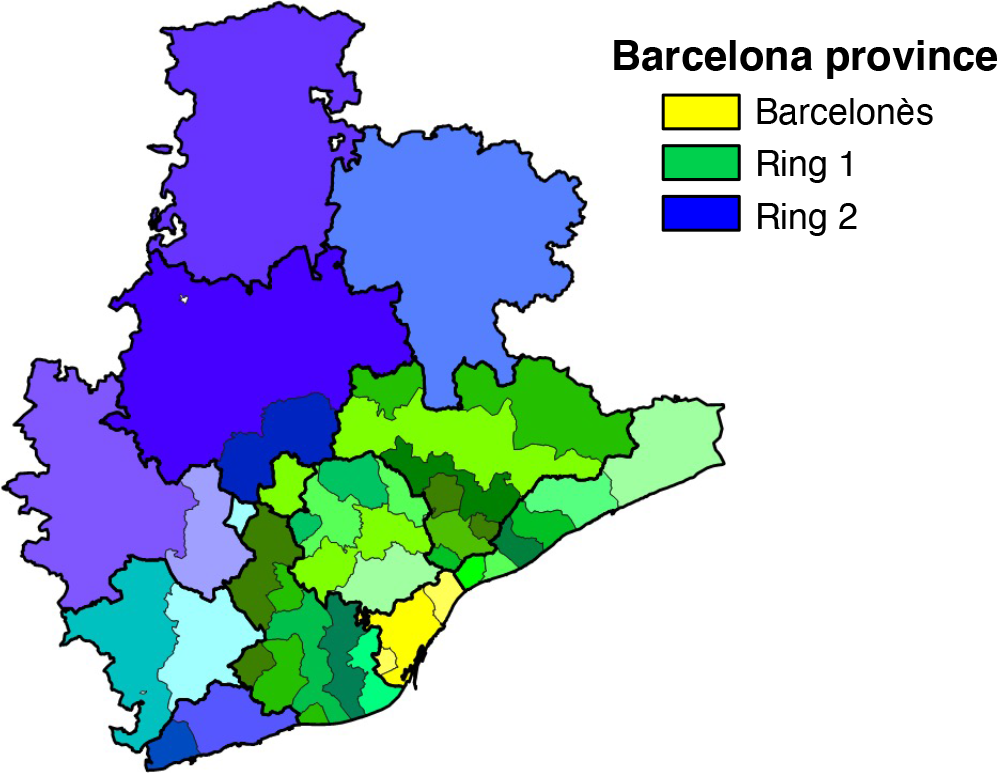
Map of the province of Barcelona. The regions used in the simulation regrouped in three concentric areas.

**Table 11:** Population in the regions defined in the province of Barcelona census file according to the county they belong to and their distance to the city of Barcelona.

| Comarca | Distance (km) | Population |
| --- | --- | --- |
| Alt Penedès | 30 – 40 | 72,309 |
|  | > 40 | 25,692 |
| Anoia | 30 – 40 | 29,429 |
|  | > 40 | 88,592 |
| Bages | 30 – 40 | 22,174 |
|  | > 40 | 161,985 |
| Baix Llobregat | < 10 | 242,777 |
|  | 10 – 15 | 253,968 |
|  | 15 – 20 | 118,991 |
|  | 20 – 25 | 111,045 |
| Baix Llobregat / Alt Penedès | 25 – 30 | 74,370 |
| Barcelonès (excluding the city of Barcelona) | < 10 | 627,672 |
| Berguedà | > 65 | 40,292 |
| Garraf | 30 – 40 | 64,702 |
|  | > 40 | 79,947 |
| Maresme | 10 – 15 | 28,159 |
|  | 15 – 20 | 66,158 |
|  | 20 – 25 | 40,793 |
|  | 25 – 30 | 134,370 |
|  | 30 – 40 | 46,859 |
|  | > 40 | 115,155 |
| Osona | > 40 | 151,652 |
| Vallès Occidental | 10 – 15 | 258,591 |
|  | 15 – 20 | 344,845 |
|  | 20 – 25 | 238,001 |
|  | 25 – 30 | 39,125 |
| Vallès Occidental / Oriental | 30 – 40 | 106,489 |
| Vallès Oriental | 10 – 15 | 21,748 |
|  | 15 – 20 | 103,441 |
|  | 20 – 25 | 25,945 |
|  | 25 – 30 | 109,488 |
|  | > 40 | 41,054 |

**Table 12:**
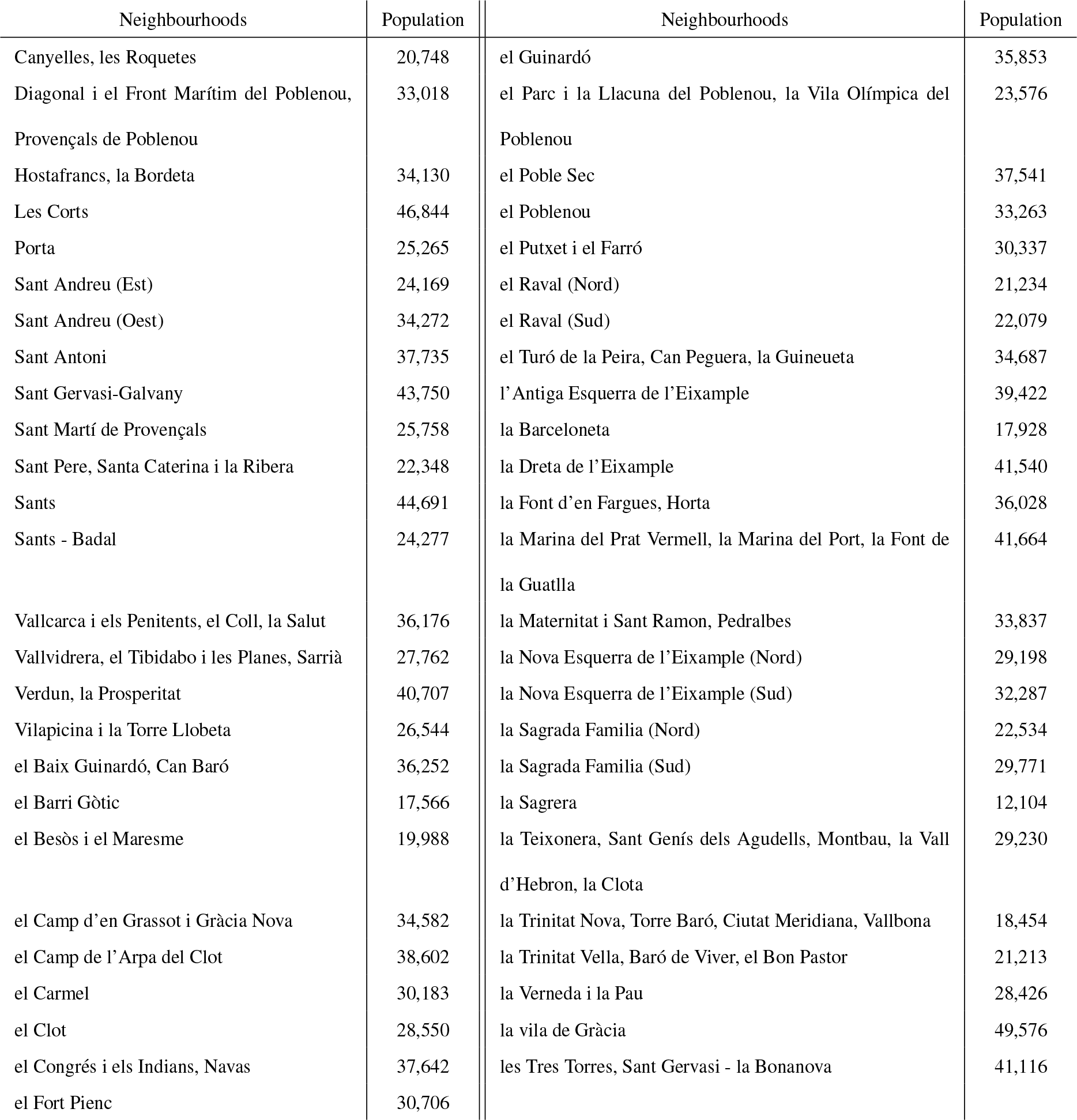
Population of the neighbourhoods of the city of Barcelona.

### Age distribution of various sectors of the population

**Figure 2:**
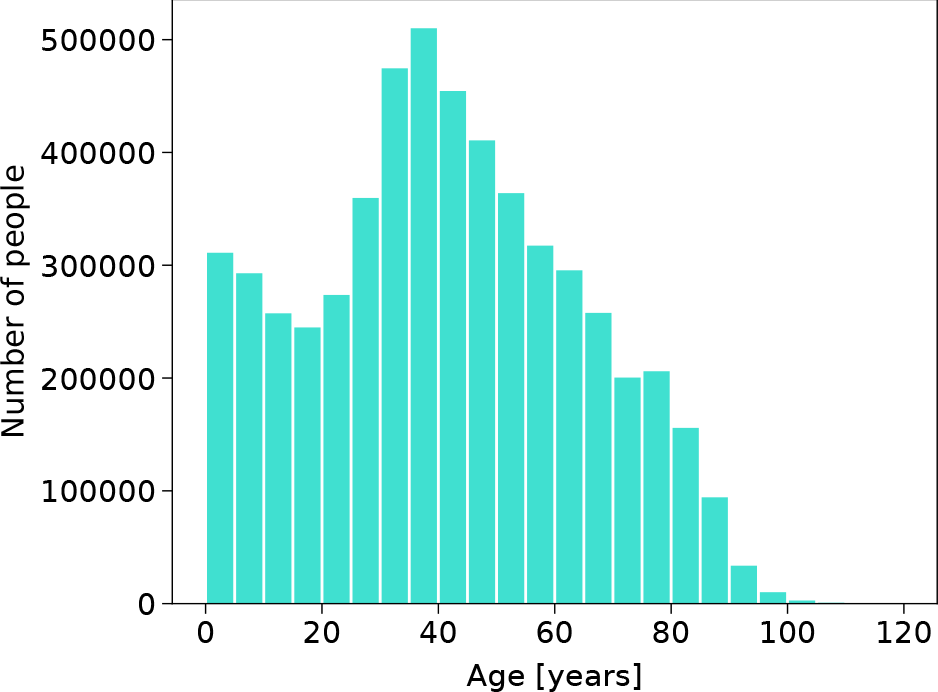
Population age. The age of the population of the province of Barcelona (5.5 million inhabitants) is shown.

**Figure 3:**
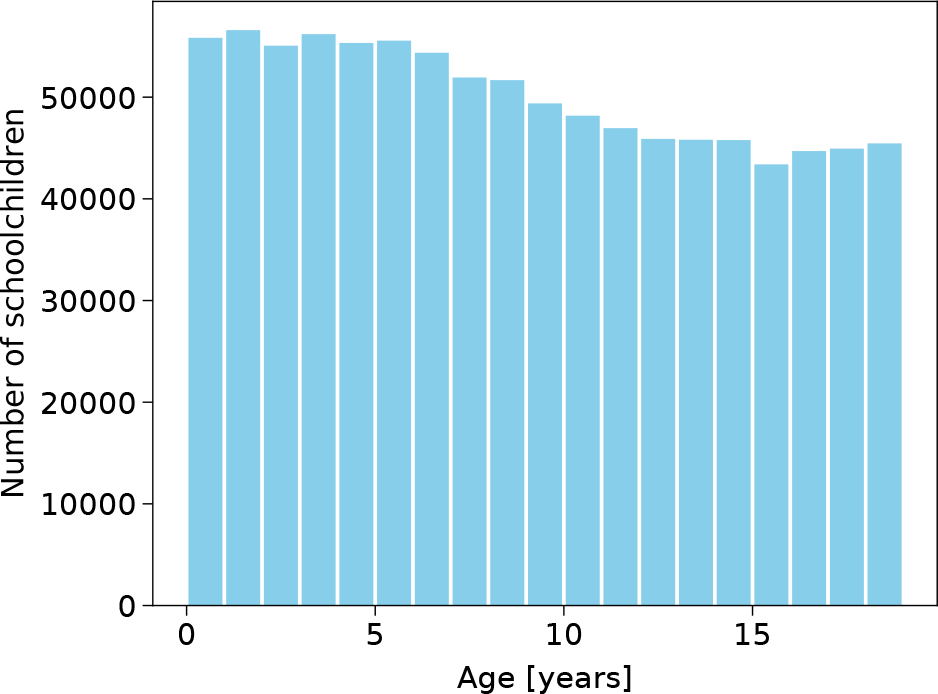
Schoolchildren age. The age of the 0 to 18-year-old children who attend school is shown.

**Figure 4:**
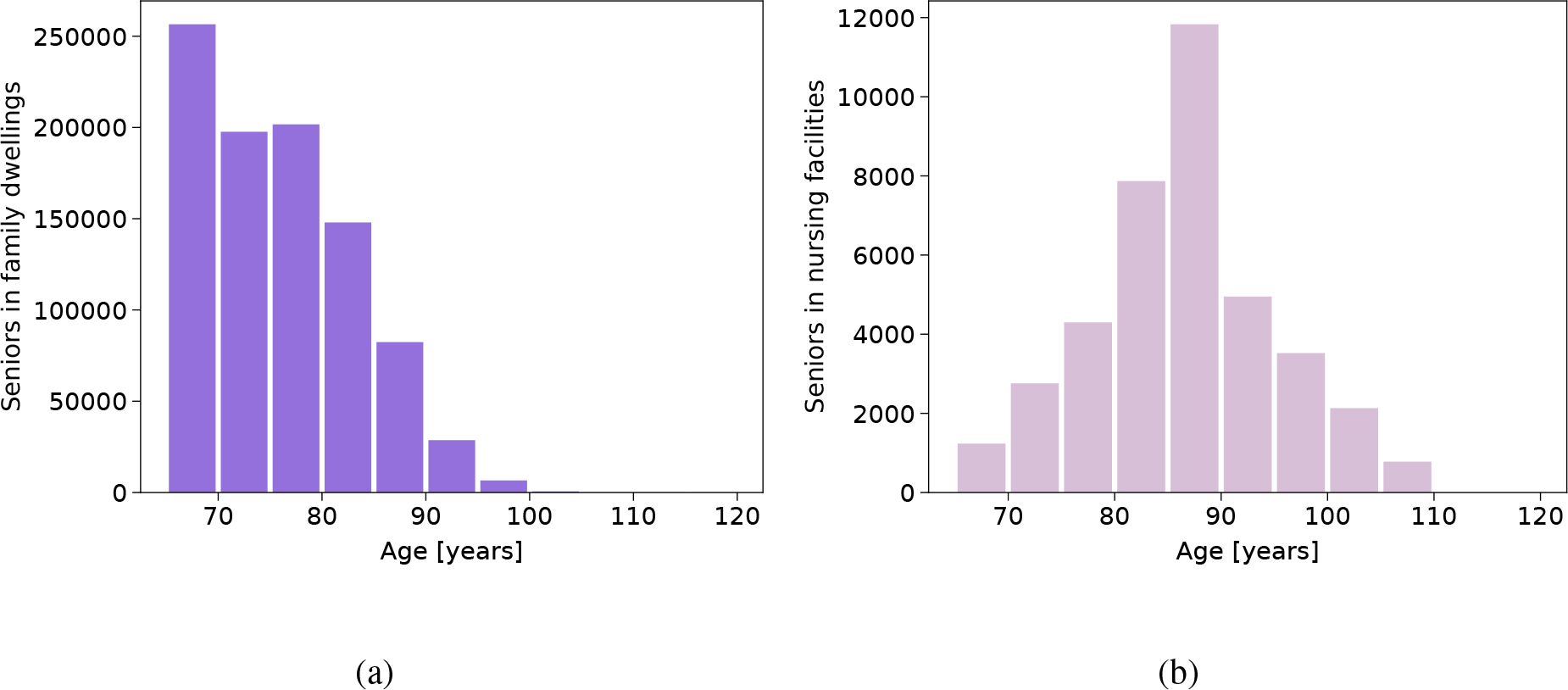
Seniors age. The age of people over 64 years of age is shown separately for **a** those who live in family dwellings, and **b** those who live in nursing facilities.

### Time profile of incubation time and viral shedding

**Figure 5:**
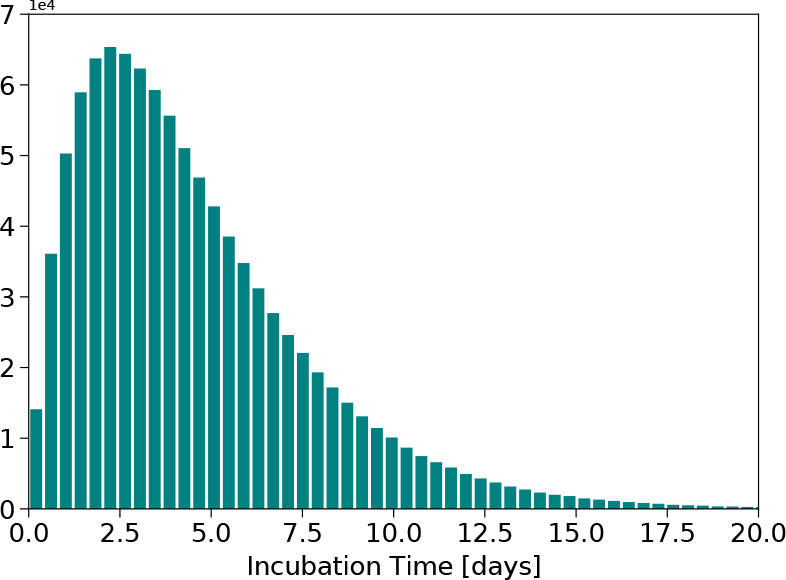
Incubation time. The time of incubation of the virus (in days) follows a Gamma distribution of mean 4.58 days and standard deviation 3.24 days.

**Figure 6:**
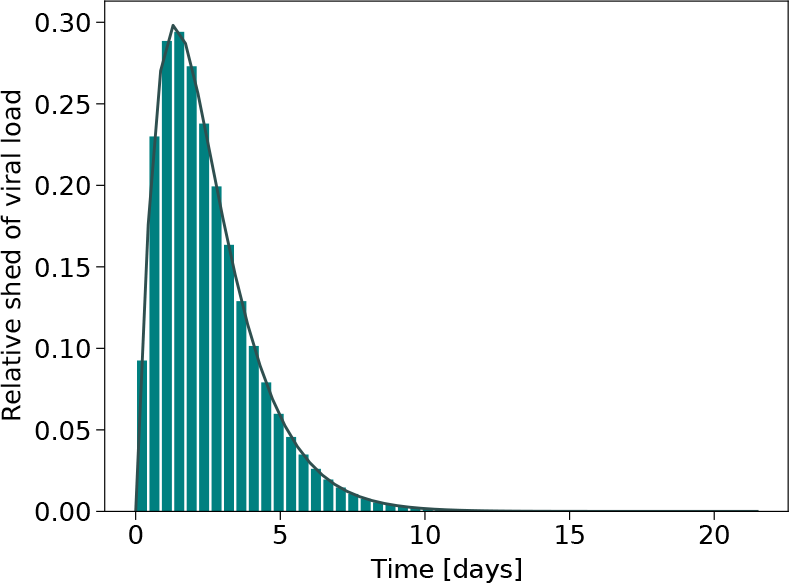
Relative profile of viral shedding. The distribution of the relative profile of the viral shedding follows a Gamma distribution of mean 2.5 days and standard deviation 1.7 days. In the simulation the distribution is truncated at 14 days.

